# Is my Arm moving? Proprioceptive impairment in Developmental Dyslexia

**DOI:** 10.1101/2020.07.30.20163923

**Authors:** Laprevotte Julie, Papaxanthis Charalambos, Saltarelli Sophie, Quercia Patrick, Gaveau Jeremie

## Abstract

Developmental Dyslexia is a specific learning disorder causing reading deficits. Although it has long been considered a purely cognitive disorder, mounting evidence revealed that Developmental Dyslexia is associated with sensory impairments. Because these impairments are restrained to vision and hearing – both senses being heavily involved in reading – a large controversy exists regarding their role in the pathophysiology of Developmental Dyslexia. Cognitive theories argue that sensory impairments are caused by the lack of reading practice, and mainly represent an aggravating factor. Sensory theories argue that damaged neural mechanisms cause sensory impairments that, themselves, cause reading disabilities. An important prediction from sensory theories is that sensory troubles should encompass the whole sensory system. Here, we directly tested proprioceptive acuity in dyslexic children and age-matched controls. We used a well-known speed perception task where a robotic manipulandum passively rotates a child’s elbow and the child presses a trigger as soon as he felts the motion. Although dyslexics and controls equally well detected salient stimuli, dyslexics were strongly impaired at detecting weaker stimuli. Furthermore, we found that proprioceptive acuity positively correlated with reading abilities. These results cannot be explained by a lack of reading practice and thus strongly support sensory theories of Developmental Dyslexia.

## Introduction

Developmental Dyslexia (DD) is a specific learning disorder causing persistent failure to acquire efficient reading. It is the most prevalent neurodevelopmental disorder, affecting about 9% of children (Katusic *et al*., 2001), and it has profound lifelong consequences on academic and professional success (Boetsch *et al*., 1996). Understanding the pathophysiological mechanisms of DD is paramount to advance this society’s grand challenge.

Thousands of studies have investigated the pathophysiology of DD, thereby documenting various impairments and developing highly debated theories (Démonet *et al*., 2004; Nicolson and Fawcett, 2019). Although DD has long been considered a purely cognitive disorder, mounting evidence revealed that DD is associated with sensory impairments (Lovegrove *et al*., 1980; Kraus *et al*., 1996; for a review see Stein, 2019). Yet, an outstanding question remains. Do sensory impairments cause DD? On one side, sensory theories argue that damaged neural mechanisms cause sensory deficiencies that, in turn, cause reading disabilities (Nicolson and Fawcett, 2019; Stein, 2019). On the other side, cognitive theories argue that this is the lack of reading practice itself that causes sensory deficits (Ramus, 2003; Goswami, 2015). Because DD is currently associated with visual and hearing impairments only – two senses that are heavily involved in reading – this long-standing controversy is still unsolved.

According to sensory theories, troubles are supposed to encompass the whole sensory system, not only vision and hearing (Nicolson and Fawcett, 2019; Stein, 2019). Testing a sensory function that is independent of reading practice would thus significantly advance the pathophysiology of DD. A positive result would undeniably support sensory theories, and vice-versa. Proprioception is an essential component of sensorimotor control (Riemann and Lephart, 2002; Proske and Gandevia, 2018), and several studies have reported sensorimotor impairments in DD (Stoodley *et al*., 2005; Pozzo *et al*., 2006). Here, while controlling for comorbidities, we directly test proprioceptive acuity in dyslexic children and age-matched controls.

## Material and Methods

### Participants

Eighteen children with developmental dyslexia (7 girls; mean age: 10.92 ± 0.78 years) and seventeen age-matched typical readers (9 girls; mean age: 11.11 ± 0.85 years) participated in this study. All were native French speakers, right-handed (Edinburgh Handedness Inventory; Oldfield, 1971), and normally integrated the school grade corresponding to their age (neither skipped nor doubled). They had normal or corrected-to-normal vision, normal hearing, and no history of neurological, proprioceptive nor psychiatric disorders. Children and parents gave their free and informed consent, and an ethics committee approved all experimental procedures (CPP Ile de France VII, France; 2017-A01547-46).

We recruited dyslexics via independent speech and occupational therapists. The diagnosis of developmental dyslexia was established by specialists (neuropsychologists, neuropediatricians, or speech therapists), using both inventories and testing procedures following the guidelines of DSM-IV or DSM-5. We recruited control children in municipal associations and schools. Neither control children nor their close family members had ever seen a medical doctor or related therapist for learning disorders.

### Clinical evaluations

We screened all children for attentional (Conners, 2008) and motor abilities (Marquet-Doléac *et al*., 2016). The aim was to exclude children at risk of Attention Deficit Hyperactivity Disorder, and/or Developmental Coordination Disorder. We excluded one dyslexic boy from the study because of his positive result at the Conners test.

A speech therapist blindly evaluated reading abilities of all children with three clinical tests: i) the Alouette Reading Test (Lefavrais, 2005), which is the French standardized assessment of reading skills; ii) the “Phonological Process” part of NEPSY 2 (Korkman *et al*., 2012), evaluating phonological skills; iii) the Timé 3 (Ecalle, 2006), that evaluates words identification. As expected, the clinical results significantly differed between the two groups (Table 1). We performed inclusion procedures and all above-mentioned clinical evaluations in one session, and the experimental sensory testing described here-after in a second session. For each child, no more than three months separated these sessions.

**Table 1.** Clinical Scores of Dyslexic and Control children at the Alouette, Nepsy2 and Timé3 tests (see methods). Efficiency Index = (A × 180) / RT, where A stands for Accuracy and is the number of words correctly read (including self-corrections), 180 is the maximum time (in seconds) allowed for the test, and RT is the reading time. Accuracy Index: (A / NWR) x 100, where A stands for Accuracy, and NWR is the number of words read.

| Test items | Dyslexics<br>N = 17 | Controls<br>N = 17 | T-test |
| --- | --- | --- | --- |
| | Mean ± SD | Mean ± SD | $t$ (34); $p$ |
| **Reading Time (s) - Alouette-R** | 178.65 ± 2.67 | 140.71 ± 26.09 | $t = 5.96$ ; **1.2e-7** |
| **Number of words read – Alouette-R** | 181.47 ± 63.29 | 258.71 ± 12.89 | $t = -4.93$ ; **2.4e-5** |
| **Number of errors – Alouette-R** | 16.82 ± 10.13 | 8.12 ± 5.01 | $t = 3.18$ ; **3.3e-3** |
| **Accuracy – Alouette-R** | 165.23 ± 65.66 | 250.41 ± 13.15 | $t = -5.24$ ; **2.8e-7** |
| **Efficiency Index – Alouette-R** | 167.12 ± 68.06 | 332.80 ± 72.64 | $t = -6.86$ ; **9.1e-9** |
| **Accuracy Index – Alouette-R** | 89.78 ± 7.50 | 96.82 ± 2.92 | $t = -3.61$ ; **1e-3** |
| **Phonological Process – NEPSY2** | 27.42 ± 12.76 | 36.23 ± 2.41 | $t = -2.79$ ; **8.7e-3** |
| **Words identification (months) – Timé3** | 29.06 ± 12.90 | -11.59 ± 21.88 | $t = 6.45$ ; **2.9e-7** |

**Table 1. Clinical Scores of Dyslexic and Control children at the Alouette, Nepsy2 and Timé3 tests (see methods).**
|  | Dyslexics<br>N = 17 | Controls<br>N = 17 | T-test |
| --- | --- | --- | --- |
| Test items | Mean ± SD | Mean ± SD | *t* (34); *p* |
| **Reading Time (s) - Alouette-R** | 178.65 ± 2.67 | 140.71 ± 26.09 | *t*= 5.96 ; **1.2e-7** |
| **Number of words read – Alouette-R** | 181.47 ± 63.29 | 258.71 ± 12.89 | *t*= -4.93 ; **2.4e-5** |
| **Number of errors – Alouette-R** | 16.82 ± 10.13 | 8.12 ± 5.01 | *t*= 3.18 ; **3.3e-3** |
| **Accuracy – Alouette-R** | 165.23 ± 65.66 | 250.41 ± 13.15 | *t*= -5.24 ; **2.8e-7** |
| **Efficiency Index – Alouette-R** | 167.12 ± 68.06 | 332.80 ± 72.64 | *t*= -6.86 ; **9.1e-9** |
| **Accuracy Index – Alouette-R** | 89.78 ± 7.50 | 96.82 ± 2.92 | *t*= -3.61 ; **1e-3** |
| **Phonological Process – NEPSY2** | 27.42 ± 12.76 | 36.23 ± 2.41 | *t*= -2.79 ; **8.7e-3** |
| **Words identification (months) – Timé3** | 29.06 ± 12.90 | -11.59 ± 21.88 | *t*= 6.45 ; **2.9e-7** |

### Sensory testing

First, to evaluate basic attention levels, children performed a classical visual and auditory simple reaction time test using salient stimuli (Lum *et al*., 2013). The order of visual and auditory tasks was pseudo-randomized. Children sat in front of a monitor with their left index finger touching the computer space key and had to press it as soon as they saw (460*550 pixels, high contrast) or heard (900 Hz, 50 dB) the stimulus. Ten visual and ten auditory stimuli were presented with a random – two to five seconds – inter-trial delay. Before the test, children performed three practice trials per sensory condition.

After a 10 minutes break, we measured proprioceptive acuity using a well-known movement detection task (Konczak *et al*., 2007; Li *et al*., 2015), which evaluates the capacity to detect the passive motion of a body-limb at various speeds. A robotic isokinetic dynamometer delivered proprioceptive stimulations (Biodex Medical Systems®, Shirley, NY, USA). Children sat on a chair and placed their right forearm on the dynamometer’s resting device. We aligned the dynamometer’s axis of rotation with the elbow flexion/extension axis of each child. The initial position of the arm was as follows: shoulder abducted between 70° and 85°, elbow flexed at 60° (0° was elbow extension), and forearm fully pronated (Konczak *et al*., 2007; Li *et al*., 2015). Within this configuration, the dynamometer passively flexed the elbow in the horizontal plane. We instructed children to relax their muscles and to focus on detecting passive flexion. Children had to press a trigger, held in their left hand, as soon as they felt the motion. Here we call “proprioceptive reaction time” the time between the motion start and the trigger pressing. The dynamometer moved with six different angular speeds: 0.25, 0.5, 1, 5, 10 and 20°.s^-1^. A total of seventy-two trials (12 trials per speed, in random order) were completed in six blocks separated by a two minutes rest time to avoid tiredness. Before the test, children performed two practice trials at each velocity (twelve trials). A subset of participants (six dyslexics and six controls) also performed the very same proprioceptive evaluation protocol on the hip joint.

Children wore opaque goggles and noise reduction headphones (Bose, QC25) to prevent sight and hearing from influencing perceptual judgments. Because changes in muscle history can lead to errors in limb position sense, we controlled for muscle thixotropy by asking children to contract the muscles around their right elbow during one second before each passive movement (Wise *et al*., 1998; Proske and Gandevia, 2018). Children were also required not to perform any vigorous motor or cognitive activity three hours before the tests. Children were authorized to take breaks at any time to prevent fatigue during the experiment.

We recorded surface electromyography (EMG) over the biceps brachii (BB), the brachioradialis (BR), and the Triceps brachii long head (TBL) to control that muscles were relaxed during proprioceptive stimulations. We recorded EMG using silver-chloride surface electrodes (7 mm recording diameter, Ag-AgCl, 20mm inter-pole distance). The skin was shaved and cleaned with alcohol. We synchronically recorded EMG signals with torque and angle using a Biopac system (Biopac MP150, Biopac System Inc, USA, gain = 1000, band-passed 1Hz – 5kHz, sampled at 10kHz).

### Data analysis

We computed all parameters using custom MATLAB programs (MathWorks).

#### Reaction time

We computed visual and auditory reaction times as the time-interval between the presentation of the stimulus and the response of the children on the keyboard/trigger. For each child and condition, we computed the mean and the variable reaction time (standard deviation of trials).

#### Electromyography

We rectified, band-passed (30-300Hz), and integrated (300ms skipping window) EMG signals. Then, we computed the ratio of EMG before movement start (300ms) divided by maximum EMG during movement, and we used this ratio to compare muscle activation/relaxation between groups.

All variables showed normal distribution (Kolmogorov-Smirnov test) and equivalent variance (Levene test). We performed group comparisons using independent two-tails *t-tests* (visual, auditory reaction times, and clinical results). We compared proprioceptive reaction times using repeated-measure ANOVA with a *group* (Controls vs. Dyslexics) and a *speed* factor (0.25°^s-1^, 0.5°^s-1^, 1°^s-1^, 5°^s-1^, 10°^s-1^ and 20°^s-1^), and *post-hoc* analyses (Scheffe) when appropriate. We also computed Pearson’s correlation coefficients between reading abilities and proprioceptive acuity. We computed a proprioceptive acuity index as the normalized average of mean and variable reaction times (at the slowest speed). We computed a reading ability index as the normalized average of four clinical scores classically used to diagnose DD in France (see Table 1): the Alouette Speed Index, the Alouette Accuracy index, the NEPSY2 result, and the Timé 3 result. We also computed Spearman correlation coefficients between arm and hip proprioceptive indexes.

Data and MATLAB codes used for their analyses are accessible upon request to the corresponding author.

## Results

### Visual and auditory simple reaction times in dyslexic and control children

To compare basic attention levels between groups, we measured simple reaction times to salient visual and auditory stimulus. Fig. 1 reveals similar attention levels in dyslexics and controls for both sensory modalities (visual: t(32) = 0.26, p = 0.79, *d* = 0.09; auditory: t(32) = 1.22, *p* = 0.23, *d* = 0.42). Since Developmental Dyslexia is associated to noisier behaviors (Sperling *et al*., 2005; Pozzo *et al*., 2006), we also compared variable reaction times – i.e., intra-participants variability – between Controls and Dyslexics. Variable visual and auditory reaction times were similar between groups (visual: t(32) = 0.19, p = 0.85, *d* = 0.19; auditory: t(32) = −0.43, *p* = 0.67, *d* = 0.15). These results demonstrate that Controls and Dyslexics were equally attentive to simple salient visual and auditory stimulations.

**Fig. 1.**
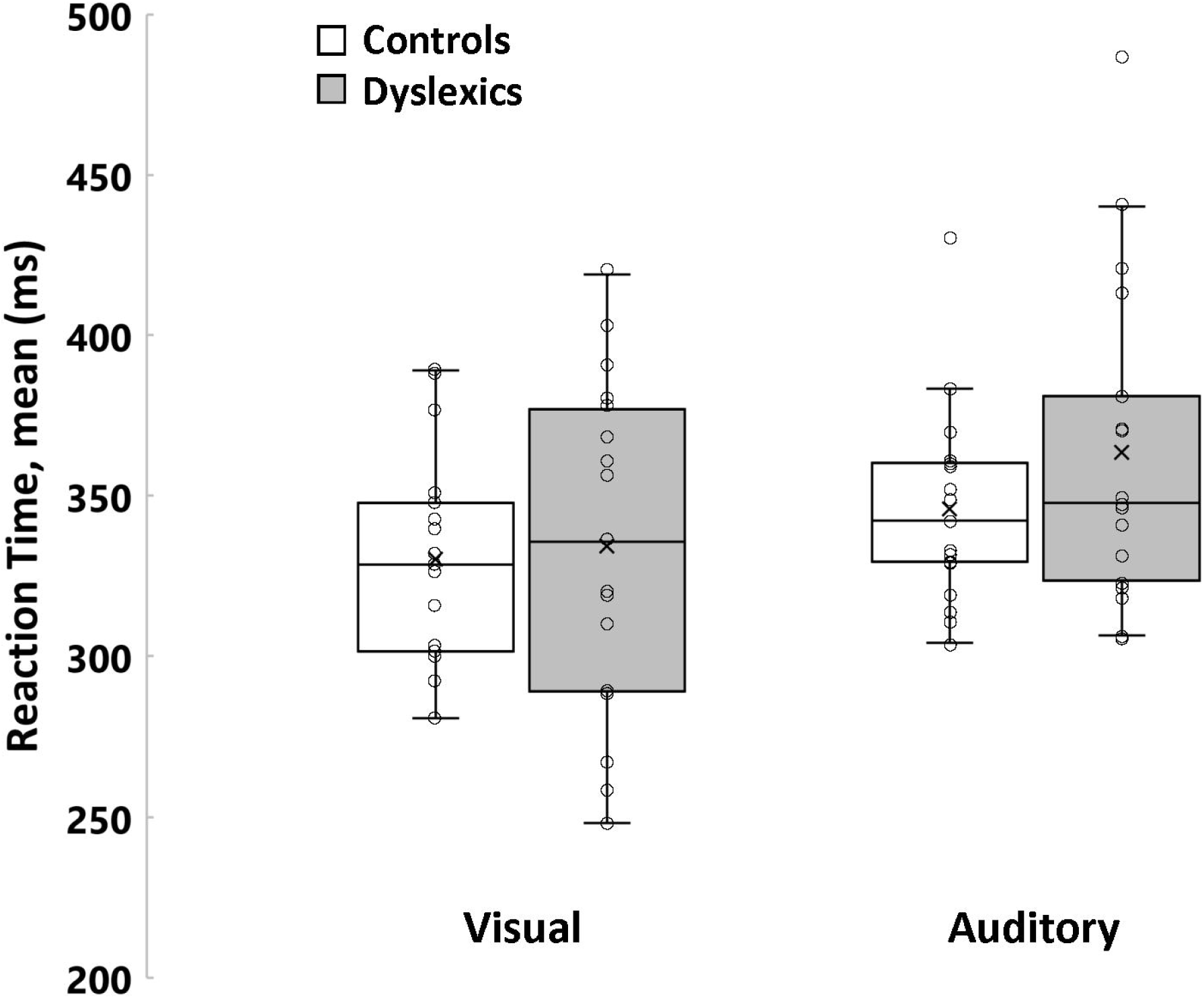
Mean Visual and Auditory Reaction Times. The box lines indicate the 25th, 50th and 75th percentiles and the whiskers extend to the 5th and 95th percentiles. Dots display individual values (n = 17 in each group) and the cross indicates the average.

### Proprioceptive acuity in dyslexic and control children

We also used a simple reaction time task to compare proprioceptive acuity between dyslexics and controls. A robotic manipulandum passively rotated their elbow joint, and children pressed a trigger as soon as they felt the motion. The robot speed changed from trial to trial, thereby modulating the proprioceptive stimulation intensity; i.e., the signal to noise ratio. Proprioceptive impairments are known to cause longer and more variable reaction times for the weakest stimulations (Konczak *et al*., 2007).

Fig. 2 (A-B) displays mean and variable proprioceptive detection times of the two groups, for the six passive movement speeds. Both Dyslexics and controls showed the well-known effect of stimulus intensity, i.e., mean and variable proprioceptive reaction times decreased when passive movement speeds increased (Konczak *et al*., 2007; Li *et al*., 2015). It is striking, however, that the slope of these relationships differed between groups. Although dyslexics and controls exhibited similar values at higher speeds, dyslexics were slower and more variable than controls at slower speeds. At the slowest speed (0.25°.s^-1^), dyslexics were even twice as long and twice as variable as controls. ANOVA yielded strong *group* x *speed* interaction effects on mean [F(5,160) 9.60; *p* < 1e-6; η^2^ = 0.23] and on variable proprioceptive reaction times [F(5,160) 7.22; *p* = 4e-6; η^2^ = 0.18]. *Post-hoc* comparisons confirmed that dyslexics were longer (*p* = 2.7e-5, d = 1.14) and more variable (*p* = 1.6e-4, d = 0.92) than controls at the slowest speed (0.25° s^-1^). This echoes previous results showing noise-related sensorimotor impairments in DD (Sperling *et al*., 2005; for a review see Nicolson and Fawcett, 2019).

**Fig. 2.**
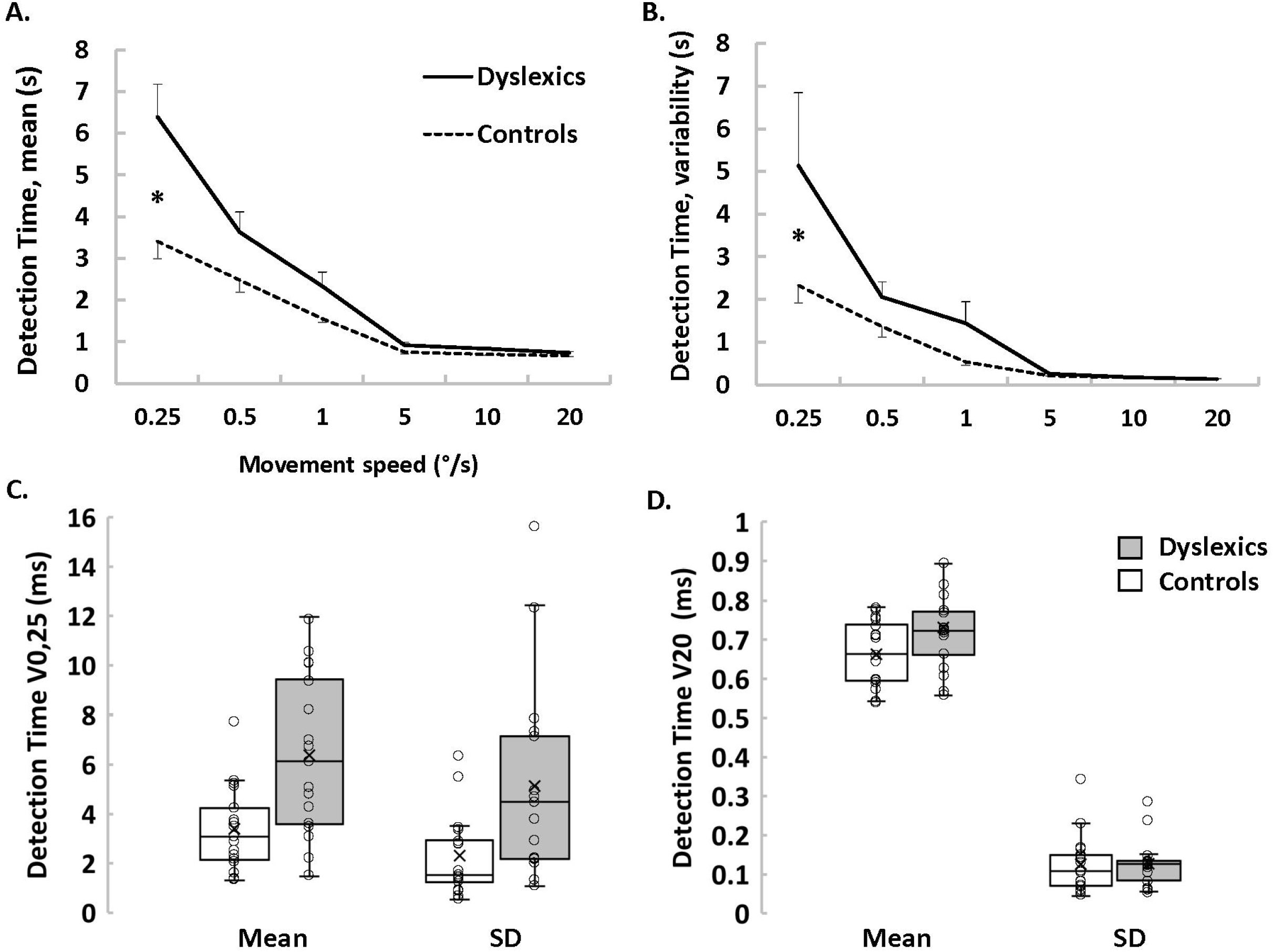
Proprioceptive detection times of passive motion at all tested speeds. (A) Mean detection time (±SE) for both groups. (B) Mean intra-participant variability of detection time (±SE) for both groups. (C) Box plots of mean and variable detection times at the slowest speed. The box lines indicate the 25th, 50th and 75th percentiles and the whiskers extend to the 5th and 95th percentiles. Dots display individual values (n = 17 in each group) and the cross indicates the average. (D) Box plots of mean and variable detection times at the highest speed. Same organization as panel C.

Box-plots in Fig. 2C-D detail mean and variable reaction times for the slowest and the highest speed. Although results obtained at the slowest speed show obvious group effects (Fig. 2C), results obtained at the highest speed demonstrate that dyslexics and controls were equally attentive to simple proprioceptive stimulations (Fig. 2D). Furthermore, comparing the first three to the last three trials did not reveal any effect in dyslexics (t(16) = 9e-3; p = 0.99, *d* = 0.04) nor in controls (t(16) = 1.16; p = 0.26, *d* = 0.01). Thus, in both groups, attention to proprioceptive stimuli was stable throughout the experiment.

To test whether the use of different coping strategies between groups could bias our proprioceptive measurements, we measured EMG activations of three major muscles around the elbow joint. T-test comparisons revealed no group effect (t(32) < 1.46, p > 0.15, d < 0.5), ensuring that dyslexics and controls equally-well respected the instruction to keep their muscles relaxed during robotic manipulations.

### Correlations between proprioception acuity and reading abilities

These results demonstrate a marked proprioceptive impairment in dyslexic children compared to age-matched controls. Next, one wonders whether a dependency relationship could exist between proprioception and dyslexia. We computed Pearson’s correlation coefficients between a proprioceptive acuity index and a reading ability index (see methods). Within these indexes, higher values mean better function.

Fig. 3 reveals a strong positive correlation between proprioceptive acuity and reading ability (Pearson R = 0.61, p = 1.3e-4). Importantly, this is not the result of a bimodal distribution of dyslexics’ and controls’ data. When pooled across groups, both indexes showed normal distribution (p > 0.2). A significant correlation existed in the dyslexic group (Pearson R = 0.52, p = 0.03) but not in the control group (Pearson R = 0.15, p = 0.57). Thus, proprioception and reading abilities are tightly linked.

**Fig. 3.**
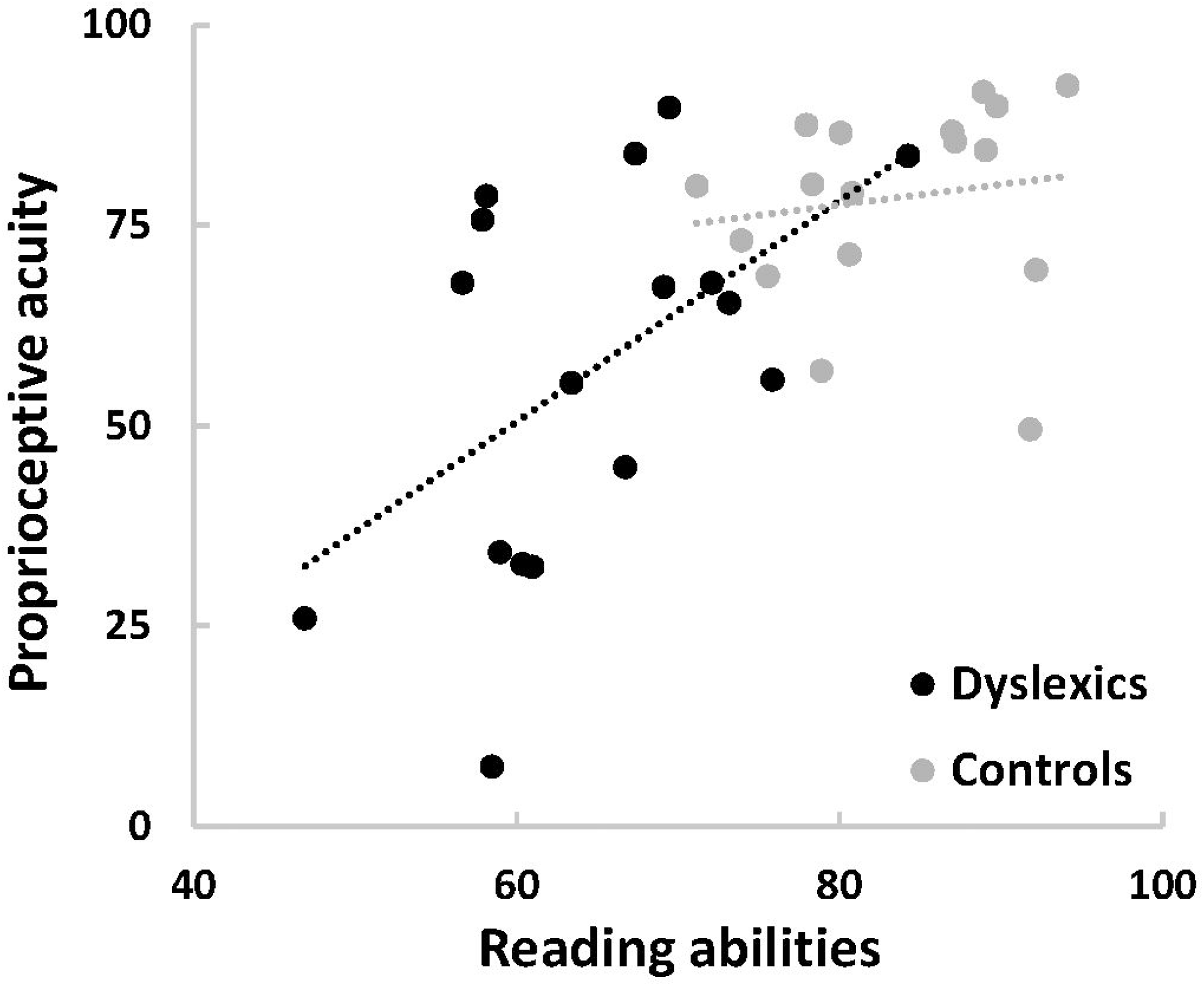
Correlation between proprioceptive acuity and reading abilities (n = 17 in each group). An index representing proprioceptive acuity (vertical axis) was computed as the normalized average of Mean and variable detection time at the slowest speed. An index representing reading abilities (horizontal axis) was computed as the normalized average of four clinical scores that are routinely used to diagnose Developmental Dyslexia (see Methods). Dashed lines depict linear regressions for each group.

Furthermore, proprioceptive indexes of the elbow and hip joints – tested in a subset of 12 participants (6 dyslexics and 6 controls) – were significantly correlated in dyslexics (Spearman R = 0.83, p = 0.04) but not in controls (Spearman R = 0,03, p = 0.96). Thus, proprioceptive impairment in dyslexia likely generalizes to other joints.

## Discussion

Our results reveal a strong and specific proprioceptive impairment in DD. Mean detection time and its variability were 100% larger in dyslexic children than in controls, and individual values correlated with clinical scores of reading abilities.

The first level of interpretation of the current results concerns the understanding of the pathophysiological mechanisms of DD. Until now, sensory impairments in DD were solely documented for vision and hearing (Lovegrove *et al*., 1980; Kraus *et al*., 1996; for a review see Stein, 2019), making them prone to the criticism that they could result from a lack of reading practice (Goswami, 2015). Because reading does not solicit arm and hip proprioception, the lack of reading practice cannot explain the large proprioceptive impairment revealed in this study. Thus, our results undeniably support the general hypothesis that damaged neural mechanisms cause widespread sensory troubles in DD (Nicolson and Fawcett, 2019; Stein, 2019). Research on other cognitive pathologies has revealed that testing sensorimotor functions allows more precocious diagnoses than testing cognitive functions (Teitelbaum *et al*., 1998; Albers *et al*., 2015). Developmental studies of sensory and sensorimotor functions in DD have the potential to further our understanding of its etiology and to develop innovative remediations (Goswami, 2015; Nicolson and Fawcett, 2019).

The second level of interpretation concerns the role played by proprioceptive impairments in DD. One may wonder whether it could directly participate in DD or instead only represent a side-effect of a widespread neural impairment. The motor theory of speech perception (Liberman and Mattingly, 1985) postulates that gesture recognition participates in the perception of phonological units. Mirror neurons allow such gesture recognition (Fadiga *et al*., 1995; Gallese *et al*., 1996), and The McGurk effect demonstrates that gesture recognition – involving proprioceptive and tactile information – indeed participates to phonological perception (Moody, 1998; Ito *et al*., 2009). In addition to vision and hearing, future work on the pathophysiology of DD shall also consider proprioception.

## Acknowledgments

The authors thank all children and their families for their participation; P. Trouilloud and all medical staff for their help with children screening and inclusion; C. Sirandre for technical help; N. Gueugneau for helpful comments on the manuscript.

## Bibliography

Albers MW, Gilmore GC, Kaye J, Murphy C, Wingfield A, Bennett DA, et al. At the interface of sensory and motor dysfunctions and Alzheimer’s disease. Alzheimer’s Dement 2015; 11: 70–98.

Boetsch EA, Green PA, Pennington BF. Psychosocial correlates of dyslexia across the life span. Dev Psychopathol 1996; 8: 539–62.

Conners K. Conners Rating Scales, manual,3rd Edition. 2008

Démonet J-F, Taylor MJ, Chaix Y. Developmental Dyslexia. Lancet 2004; 363: 1451–60.

Ecalle J. Timé 3, test d’identification de mots écrits pour enfants de 7 à 15 ans. Mot à Mot. 2006

Fadiga L, Fogassi L, Pavesi G, Rizzolatti G. Motor facilitation during action observation: A magnetic stimulation study. J Neurophysiol 1995; 73: 2608–11.

Gallese V, Fadiga L, Fogassi L, Rizzolatti G. Action recognition in the premotor cortex. Brain 1996; 119: 593–609.

Goswami U. Sensory theories of developmental dyslexia: Three challenges for research. Nat Rev Neurosci 2015; 16: 43–54.

Ito T, Tiede M, Ostry DJ. Somatosensory function in speech perception. Proc Natl Acad Sci U S A 2009; 106: 1245–8.

Katusic SK, Colligan RC, Barbaresi WJ, Schaid DJ, Jacobsen SJ. Incidence of reading disability in a population-based birth cohort, 1976-1982, Rochester, Minn. Mayo Clin Proc 2001; 76: 1081–92.

Konczak J, Krawczewski K, Tuite P, Maschke M. The perception of passive motion in Parkinson’s disease. J Neurol 2007; 254: 655–63.

Korkman M, Kirk U, Kemp S. NEPSY-II, Bilan Neuropsychologique de l’enfant, 2de édition. ECPA. 2012

Kraus N, McGee TJ, Carrell TD, Zecker SG, Nicol TG, Koch DB. Auditory neurophysiologic responses and discrimination deficits in children with learning problems. Science (80-) 1996; 273: 971–3.

Lefavrais P. Alouette-R, Test d’analyse de la vitesse en lecture à partir d’un texte. Forme révisée. Editions d. 2005

Li K-Y, Su W-J, Fu H-W, Pickett KA. Kinesthetic deficit in children with developmental coordination disorder. Res Dev Disabil 2015; 38C: 125–33.

Liberman AM, Mattingly TH. The motor theory of speech perception revisited. Cognition 1985; 21: 1–36.

Lovegrove WJ, Bowling A, Badcock D, Blackwood M. Specific Reading Disability?: Differences in Contrast Sensitivity as a Function of Spatial Frequency. Science 1980; 210 (4468): 439–40.

Lum JAG, Ullman MT, Conti-Ramsden G. Procedural learning is impaired in dyslexia: Evidence from a meta-analysis of serial reaction time studies. Res Dev Disabil 2013; 34: 3460–76.

Marquet-Doléac J, Soppelsa R, Albaret J. MABC-2 - Batterie d’évaluation du mouvement chez l’enfant - 2nde édition. Adaptation Française. Pearson; 2016

Moody J. Proprioception and the McGurk Effect. Science (80-) 1998; 282: 1991.

Nicolson RI, Fawcett AJ. Development of Dyslexia: The Delayed Neural Commitment Framework. Front Behav Neurosci 2019; 13: 1–16.

Pozzo T, Vernet P, Creuzot-Garcher C, Robichon F, Bron A, Quercia P. Static postural control in children with developmental dyslexia. Neurosci Lett 2006; 403: 211–5.

Proske U, Gandevia SC. Kinesthetic senses. Compr Physiol 2018; 8: 1157–83.

Ramus F. Developmental dyslexia: specific phonological deficit or general sensorimotor dysfunction? Neurobiology 2003; 13: 212–8.

Riemann BL, Lephart SM. The sensorimotor system, Part II: The role of proprioception in motor control and functional joint stability. J Athl Train 2002; 37: 80–4.

Sperling AJ, Lu ZL, Manis FR, Seidenberg MS. Deficits in perceptual noise exclusion in developmental dyslexia. Nat Neurosci 2005; 8: 862–3.

Stein J. The current status of the magnocellular theory of developmental dyslexia. Neuropsychologia 2019; 130: 66–77.

Stoodley CJ, Fawcett AJ, Nicolson RI, Stein JF. Impaired balancing ability in dyslexic children. Exp brain Res 2005; 167: 370–80.

Teitelbaum P, Teitelbaum O, Nye J, Fryman J, Maurer RG. Movement analysis in infancy may be useful for early diagnosis of autism. Proc Natl Acad Sci U S A 1998; 95: 13982–7.

Wise AK, Gregory JE, Proske U. Detection of movements of the human forearm during and after co-contraction of muscle acting at the elbow joint. J Physiol 1998; 508: 325–30.

